# Observer agreement and clinical significance of chest CT reporting in patients suspected of COVID-19

**DOI:** 10.1101/2020.05.07.20094102

**Authors:** Marie-Pierre Debray, Helena Tarabay, Lisa Males, Nisrine Chalhoub, Elyas Mahdjoub, Thomas Pavlovsky, Benoît Visseaux, Donia Bouzid, Raphael Borie, Catherine Wackenheim, Bruno Crestani, Christophe Rioux, Loukbi Saker, Christophe Choquet, Jimmy Mullaert, Antoine Khalil

## Abstract

**Objectives:** To assess inter-observer agreement and clinical significance of chest CT reporting in patients suspected of COVID-19.

**Methods:** From 16th to 24th March 2020, 241 consecutive patients addressed to hospital for COVID-19 suspicion had both chest CT and SARS-CoV-2 RT-PCR. Eight observers (2 thoracic and 2 general senior radiologists, 2 junior radiologists and 2 emergency physicians) retrospectively categorized each CT into one out of 3 categories (evocative, compatible for COVID-19 pneumonia, and not evocative or normal). Observer agreement for categorization between all readers and pairs of readers with similar experience was evaluated with the Kappa coefficient. The results of a consensus categorization were correlated to RT-PCR.

**Results:** Observer agreement across the 3 categories was good between all readers (κ value 0.68 95%CI 0.67-0.70) and good to very good between pairs of readers (0.64-0.85). It was very good (κ 0.81 95%CI 0.79-0.83), fair (κ 0.32 95%CI 0.29-0.34) and good (κ 0.74 95%CI 0.71-0.76) for the categories evocative, compatible and not evocative or normal, respectively. RT-PCR was positive in 97%, 50% and 27% of cases classified in the respective categories. Observer agreement was lower (p=0.045) and RT-PCR positive cases were less frequently categorized evocative in presence of an underlying pulmonary disease (*p*<0.001).

**Conclusion:** Inter-observer agreement for chest CT reporting using categorization of findings is good in patients suspected of COVID-19. Among patients considered for hospitalization in an epidemic context, CT categorized evocative is highly predictive of COVID-19, whereas the predictive value of CT decreases between the categories compatible and not evocative.

**Key results:**

1. Inter-observer agreement for chest CT reporting into categories is good in patients suspected of COVID-19
2. Chest CT can participate in estimating the likelihood of COVID-19 in patients presenting to hospital during the outbreak, CT categorized «evocative» being highly predictive of the disease whereas up to a quarter of patients with CT «not evocative» had a positive RT-PCR in our study.
3. Observer agreement is lower and CTs of positive RT-PCR cases less frequently “evocative” in presence of an underlying pulmonary disease

## Introduction

Since December 2019, a new respiratory disease related to a new coronavirus, SARS-CoV-2, developed in China and rapidly spread to other countries, reaching a pandemic stage in March 2020 [1,2]. Even if the disease follows a benign course in many cases, some patients develop respiratory difficulties requiring hospitalization, leading to a large amount of patients with clinical suspicion of coronavirus disease 2019 (COVID-19) presenting to the emergency departments [3]. Accurate identification of COVID-19 patients is crucial to isolate them from not infected patients and to limit the diffusion of the outbreak. The reference standard is the positivity of the real-time reverse transcription-polymerase chain reaction (RT-PCR); nevertheless, the sensitivity of this test remains unclear, having been reported between 42 and 71 % in some early series [4–6], because of suboptimal sampling technique, limitations in performance assay or low viral load in the nasopharyngeal area. Chest CT shows abnormalities in a large majority of cases, with some signs described as typical or very evocative of the disease in the current outbreak context [7–11]. Sensitivity of chest CT has been reported as high as 97% as compared to RT-PCR [4] and CT abnormalities could precede RT-PCR positivity [12]. Because of its readily availability, chest CT may assist first-line triage of patients presenting to hospital [3]. Several Radiology Societies [6,13–15] have proposed structured reporting of CT into categories, defined according to the typical or less typical appearance of lung involvement, to facilitate communication with physicians. In routine practice, categorization is based on each reader individual impression supported by numerous papers having described imaging signs of COVID-19 pneumonia [7–11]. Such categorization may directly impact the clinical decision-making. However, the reproducibility of the categorization is unknown and the clinical significance of the different categories is unclear. Thus, the objectives of our study were to assess inter-observer agreement to categorize CT findings as well as performances of chest CT across the different categories in patients suspected of COVID-19 presenting to hospital.

## Material and methods

### Setting

This is a monocentric retrospective study conducted in a University Hospital (Bichat Claude-Bernard Hospital, Paris, France) between 2020 March 16th and 2020 March 24th. Institutional review board was approved and written informed consent waived. During this period of COVID-19 outbreak, patients presenting at our hospital for COVID-19 suspicion and for whom hospitalization was considered had both chest CT-scan and SARS-CoV-2 RT-PCR. Diagnosis of COVID-19 relied on the positivity of the RT-PCR and CT could assist early triage in critically ill patients or with clinically overt pneumonia. Patients with a negative RT-PCR result could have a subsequent RT-PCR test and/or another chest CT during the next few days, depending on the physician’s judgment.

### Patients

Consecutive adult patients attending the emergency room or the infectious diseases department of our hospital with clinical suspicion of COVID-19 and having both chest CT and SARS-CoV-2 RT-PCR were included.

### Clinical and laboratory data collection

Demographic, clinical and laboratory data at presentation, follow-up data when available were extracted from electronic medical records. Clinical data included symptoms, any need for oxygen supply, time from symptom onset to CT, comorbidities and pre-existing pulmonary diseases. RT-PCR was performed on nasopharyngeal swabs or aspiration, using RealStar® SARS-CoV-2 RT-PCR Kit (Altona Diagnostics) or Cobas® SARS-CoV-2 Test (Roche).

### Chest CT protocols

Chest CT-scans were acquired on a multidetector-row CT (Canon Aquilion PRIME or GENESIS) without contrast medium injection. They were performed in the supine position at full inspiration. The scanning parameters were as follows: 120 kVp, automatic exposure control for tube current (SD:15), exposure time 0.27-0.35 sec per rotation depending on the CT unit, collimation 40 mm. Images were reconstructed with 1 mm slice thickness and 0.8 mm inter-slice gap, using a high-frequency reconstruction algorithm.

### Image analysis

All CT-scans were analyzed by 8 readers, including 2 senior emergency physicians (TP, DB, with 10 years of experience each), 2 radiology residents (HT, NC, 4 and 5 years of experience), 2 senior general radiologists (LM, EM, 6 and 9 years of experience) and 2 senior thoracic radiologists (MPD, AK, 23 and 25 years of experience), blindly to RT-PCR results and final diagnosis. All readers classified each examination into one out of 4 categories, as follows: evocative, compatible, not evocative of COVID-19 and normal following the recommendations of the French Society of Radiology [14]. The global impression of each reader was supported by previous typical and less typical reported signs in the literature. A guide was provided, recalling these signs. The “evocative” category included multifocal ground-glass opacities (GGO), being nodular or not, or crazy-paving with or without consolidations, with a bilateral, peripheral or mixed distribution and involvement of the posterior zones. The intermediate category “compatible” corresponded to cases showing abnormalities already reported in COVID-19 but that may be encountered in other diseases or very limited in extent. It included GGO and/or consolidations with very few lesions and unilateral distribution, exclusive central distribution or absence of posterior lung areas involvement, halo sign as main abnormality, and association of typical opacities, with atypical signs or other lesions. The category “not evocative” corresponded to cases showing abnormalities very rarely reported in COVID-19 or typical of another diagnosis as isolated systematized consolidation, discrete centrilobular nodules with tree-in-bud appearance or lung cavitation in favor of other lung infection, centrally distributed GGO with septal lines and pleural effusion in favor of cardiogenic pulmonary edema, peripheral reticulations with or without honeycombing, traction bronchiectasis and GGO in favor of interstitial lung disease. This category also included non-specific abnormalities as sub-segmental atelectasis or opacities considered to be sequelae. We chose to merge a normal appearance of the lung parenchyma with these non-specific signs because its distinction from minor non-specific abnormalities or typical of sequelae does not have clinical relevance in the present study. Consequently, 3 categories were retained: evocative, compatible and not evocative or normal.

Any disagreement between the 4 senior radiologists was analyzed in consensus of these 4 readers giving a final consensus categorization for all cases.

Finally, all chest CTs were described by one thoracic radiologist (MPD) for presence and distribution of various elementary signs, as well as signs of any underlying pulmonary disease (significant pulmonary emphysema, interstitial lung disease, bronchiectasis, parenchymal sequelae, bronchial carcinoma).

### Statistical analysis

Categorical variables were described by numbers and percentage for each category. The agreement between two and more than two readers was evaluated with the Cohen’s kappa coefficient and the Fleiss’ kappa, respectively, and their 95% confidence interval, which measures the excess proportion of agreement after taking chance into account.

Comparisons between dependent kappas (e.g. for different couple of readers for the same images) were performed with bootstrapping (N=10000 samples) and the *p*-value corresponds to the proportion of bootstrap samples that yield a couple of kappa value in a different order than the observed one. Comparison between independent kappa (e.g. for different levels of a categorical variable) were performed according to the method proposed in [16].

Comparison of the frequency of radiologic signs between categories was performed with the fisher exact test. All analysis were done using R v3.6.1.

## Results

### General description of the population

In total, 241 patients were included. Their demographic, signs at presentation and comorbidities are in Table 1. COVID-19 was confirmed in 158 patients by RT-PCR positivity. COVID-19 was deemed likely despite two negative RT-PCR in 2 cases with clinical and CT follow-up strongly supportive of this diagnosis. 15 patients were considered non-COVID-19 because of at least 2 consecutive negative RT-PCR and absence of clinical and radiological signs favoring COVID-19 during follow-up. 66 patients were considered non-COVID-19 with only one negative RT-PCR but including 38 with CT and/or clinical follow-up (Fig. 1).

**Figure 1.**
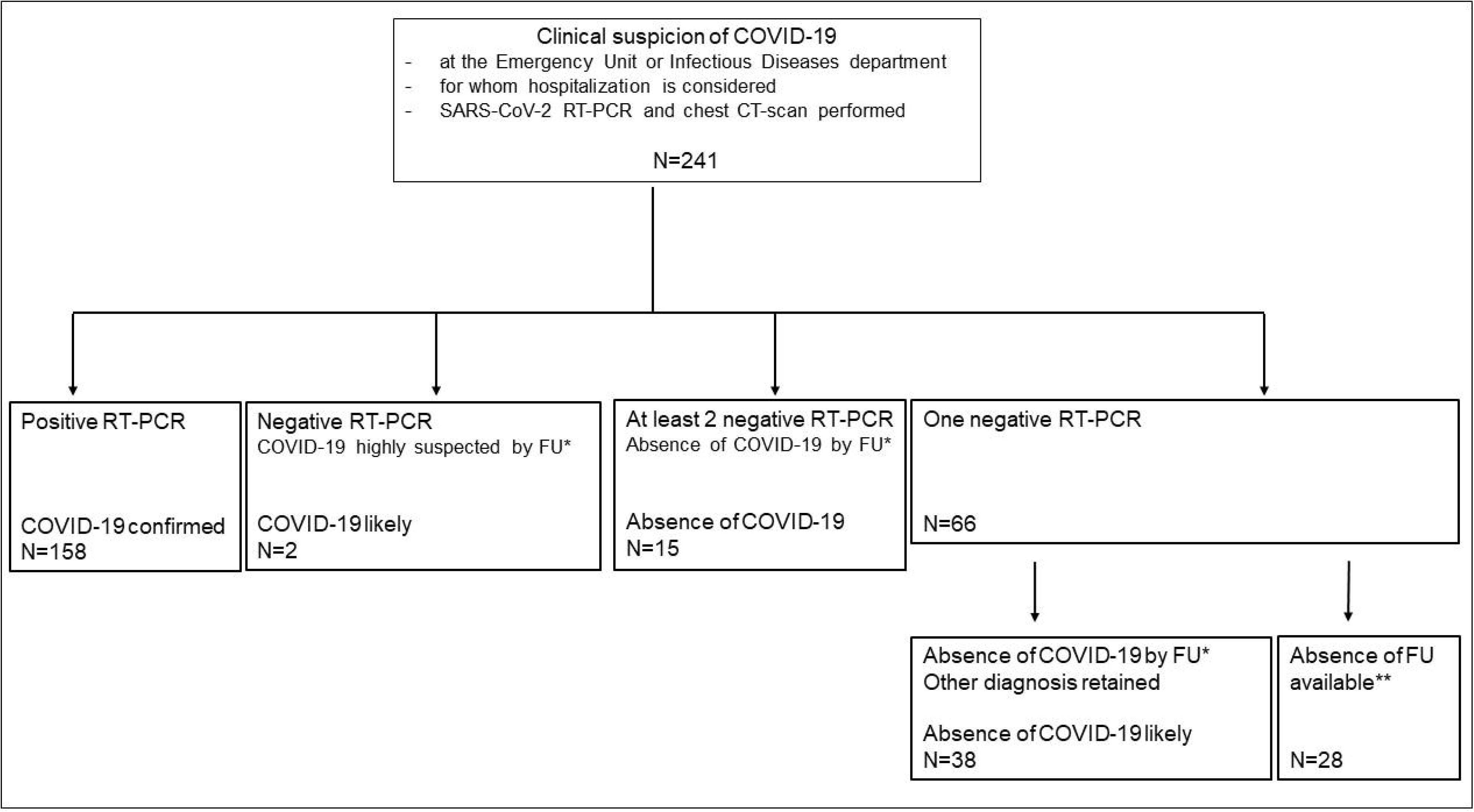
Flow chart of the study. SARS-CoV-2 RT PCR and chest CT were performed at presentation, except in 7 cases for which RT-PCR had been performed one to 3 days earlier. Among 158 COVID-19 confirmed cases, RT-PCR was positive at admission in 151 cases and in subsequent days during hospitalization in 7 cases.^*^Clinical and chest CT follow-up. ^**^Absence of follow-up available because the patient had been transferred to another hospital or turned back home.

**Table 1.** Clinical characteristics of 241 included patients with clinical suspicion of COVID-19.

| Characteristics | No.(%) or Median [Q1-Q3] |
| --- | --- |
| Age (years) | 64 [52-79] |
| Male (%) | 155 (64) |
| Comorbidities |  |
| COPD | 24 (10) |
| Respiratory insufficiency | 37 (15) |
| Any other | 170 (71) |
| Signs at presentation |  |
| Respiratory (any) | 202 (84) |
| Cough | 152 (63) |
| Expectoration | 27 (11) |
| Dyspnea | 149 (62) |
| Chest pain | 34 (14) |
| Need for Oxygen supply | 143 (59) |
| Digestive (abdominal pain, diarrhea, nausea) | 65 (27) |
| Neurologic (confusion, headache) | 53 (22) |
| Asthenia | 92 (38) |
| Fever, chills, sweats | 115 (48) |
| Onset-to-CT-delay (days) | 4 [2-7] |
| Data are numbers with percentages in brackets or medians with lower and upper quartiles in square brackets. |  |

### Inter-observer agreement for chest CT categorization

Kappa coefficient between all readers across the 3 CT categories was good (0.68, 95%CI 0.67-0.70). It was good to very good between each pair of readers, and significantly better between resident radiologists as compared to thoracic senior radiologists (*p*<0.001) (Table 2). The Kappa value between all readers was lower when abnormalities were unilateral as compared to bilateral lesions (*p*=0.018) and in presence of underlying pulmonary lesions (*p*=0.045). It was lower for patients older as compared to those younger than 70 years (*p*=0.017), and when time from symptom onset to CT was shorter than 5 days (*p*=0.012). Observer agreement was very good between all readers for the category “evocative” (0.81, 95%CI 0.79-0.83), it was good for the category “not evocative or normal” (0.74, 95%CI 0.71-0.76) and fair for the category “compatible” (0.32, 95%CI 0.29-0.34) (Table 3).

**Table 2.** Chest CT categorization of the 8 readers and consensus reading, with observer agreement between pairs and all readers.

| | Evocative<br>N (%) | Compatible<br>N (%) | Not evocative<br>or normal<br>N (%) | $\kappa$ value<sup>a</sup><br>(95% CI) |
| --- | --- | --- | --- | --- |
| Thoracic senior radiologists |  |  |  |  |
| Reader 1 | 117 (49) | 32 (13) | 92 (38) | 0.69 |
| Reader 2 | 126 (52) | 52 (22) | 63 (26) | (0.62-0.76) |
| General senior radiologists |  |  |  |  |
| Reader 3 | 119 (49) | 18 (7.5) | 104 (43) | 0.75* |
| Reader 4 | 112 (51) | 38 (11) | 91 (39) | (0.68-0.82) |
| Resident radiologists |  |  |  |  |
| Reader 5 | 122 (51) | 26 (11) | 93 (39) | 0.85** |
| Reader 6 | 114 (47) | 29 (12) | 98 (41) | (0.79-0.90) |
| Senior emergency physicians |  |  |  |  |
| Reader 7 | 119 (49) | 37 (15) | 85 (35) | 0.64* |
| Reader 8 | 131 (54) | 35 (15) | 75 (31) | (0.56-0.72) |
| All readers |  |  |  | 0.68<br>(0.67-0.70) |
| Consensus reading | 123 (51) | 30 (12) | 88 (37) |  |

Data are numbers with percentages, or 95% confidence interval for the $\kappa$ value, in brackets
<sup>a</sup>Cohen's kappa value for agreement between 2 readers, and Fleiss' kappa value for agreement between all readers
\*Observer agreement not significantly different between general senior radiologists nor senior emergency physicians and agreement between thoracic senior radiologists
\*\*Observer agreement significantly better between resident radiologists as compared to agreement between thoracic senior radiologists ( $p < 0.001$ )

### Correlation of RT-PCR and categories on chest CT

The RT-PCR positivity rate was highly significantly different among the 3 categories (*p*<0.0001): 119 out of 123 (97%), 15 out of 30 (50%) and 24 out of 88 (27%) cases considered evocative, compatible and not evocative or normal by the consensus reading had a positive RT-PCR, respectively (Table 3). The rate of RT-PCR positivity was 31% (22 out of 70) among not evocative CTs and 11% (2 out of 18) in normal CTs.

**Table 3.** Inter-observer agreement for each category and correlation to SARS-CoV-2 RT-PCR results.

|  | Evocative | Compatible | Not evocative<br>or normal | *P* value |
| --- | --- | --- | --- | --- |
| Kappa value, all readers<br>(95% CI) | 0.81<br>(0.79-0.83) | 0.32<br>(0.29-0.34) | 0.74<br>(0.71-0.76) |  |
| Effective for comparison to<br>RT-PCR (N)<br>(consensus reading) | 123 | 30 | 88 |  |
| Positivity of RT-PCR (%) | 119 (97) | 15 (50) | 24 (27) | <0.0001 |
| Data are numbers with percentages, or 95% confidence interval for the $\kappa$ value, in brackets | | | | |

With RT-PCR as reference, chest CT classified evocative had 75% sensitivity (95%CI 68-81%) and 95% specificity (95%CI 87-98%) whereas chest CT classified evocative or compatible had 85% sensitivity (95%CI 78-90%) and 77% specificity (95%CI 66-85%) for COVID-19. Sensitivity for evocative CT was significantly lower for patients with a delay since symptom onset lower than 5 days (68% 95%CI [60-78%] vs 83% 95%CI [73-91%], *p*=0.038) and for patients with an underlying pulmonary disease (36% 95%CI [21-54%] vs 87% 95%CI [80-92%], p<0.001), but the difference was not significant between younger or older patients (age>70, *p*=0.23).

Among 90 patients with a first negative RT-PCR, 22 were re-tested, including 5 out of 6 patients (83%) with CT considered evocative, 5 out of 17 patients (29%) with CT considered compatible and 12 out of 67 patients (18%) with CT considered not evocative or normal. Of these subsequent RT-PCR, 2 out of 5 were positive for each category «evocative» and «compatible» and 3 out of 12 for the third category.

### Clinical and chest CT findings in the different categories

CT features of the whole population, of RT-PCR positive cases and of the different CT categories are in Table 4. The most frequent pattern of CT considered evocative was mixed with predominant GGO. Typical bilateral and peripheral distribution with posterior involvement was almost constant. Some centrally distributed lesions were associated to peripheral lesions in 72% of cases (Fig. 2). The 30 chest CTs considered compatible more frequently showed pure GGO as compared to evocative cases (*p*=0.0012). Among these 30 cases, 12 and 6 showed features of an underlying pulmonary disease and of an associated pulmonary edema, respectively (Fig. 3, 4). As compared with cases classified «not evocative» (Fig. 5, 6), cases classified compatible more frequently showed a typical distribution and atypical signs were absent among those with positive RT-PCR. Time from symptom onset to CT was longer, patients were older and need for oxygen supply was more frequent in patients whose CT was categorized evocative as compared to other patients.

**Table 4.** CT features in all cases, SARS-CoV-2 RT-PCR positive cases and in the different CT categories

|  | All<br>N=241<br>N (%) | Positive<br>RT-PCR<br>N=158<br>N (%) | Evocative<br>N=123<br>N (%) | Compatible<br>N=30<br>N (%) | Not<br>evocative<br>N=70<br>N (%) | *P* value* |
| --- | --- | --- | --- | --- | --- | --- |
| **Pattern** |  |  |  |  |  |  |
| Isolated GGO | 51 (21) | 31 (20) | 21 (17) | 14 (47) | 16 (23) | 0.004 |
| Isolated consolidations | 24 (10) | 8 (5) | 1 (1) | 4 (13) | 19 (27) | <0.001 |
| Mixed, predominant GGO | 91 (38) | 82 (52) | 81 (66) | 7 (23) | 3 (4) | <0.001 |
| Mixed, predominant consolidations | 30 (12) | 24 (15) | 20 (16) | 4 (13) | 6 (9) | 0.33 |
| **Distribution** |  |  |  |  |  |  |
| Bilateral | 177 (73) | 140 (89) | 122 (99) | 24 (80) | 31 (44) | <0.001 |
| Posterior | 181 (75) | 139 (88) | 122 (99) | 25 (83) | 34 (49) | <0.001 |
| Peripheral | 186 (78) | 143 (91) | 122 (99) | 24 (80) | 40 (59) | <0.001 |
| Central | 145 (61) | 114 (72) | 107 (87) | 15 (50) | 23 (34) | <0.001 |
| **Atypical features** |  |  |  |  |  |  |
| Pleural effusion | 35 (15) | 13 (8) | 10 (8) | 8 (27) | 17 (24) | 0.002 |
| Systematized consolidation | 7 (3) | 4 (3) | 1 (1) | 1 (3) | 5 (7) | 0.035 |
| Centrilobular nodules | 14 (6) | 4 (3) | 1 (1) | 2 (7) | 10 (14) | <0.001 |
| Mucoid impaction | 18 (7) | 6 (4) | 2 (2) | 4 (13) | 12 (17) | <0.001 |
| Underlying pulmonary disease | 61 (25) | 36 (23) | 13 (11) | 14 (47) | 33 (47) | <0.001 |

Cases showing normal lung parenchyma are not included.
\**P* value is for comparison between CT categories
GGO: ground-glass opacities
Underlying pulmonary disease includes significant emphysema, interstitial lung disease, bronchiectasis, sequelae

**Figure 2.**
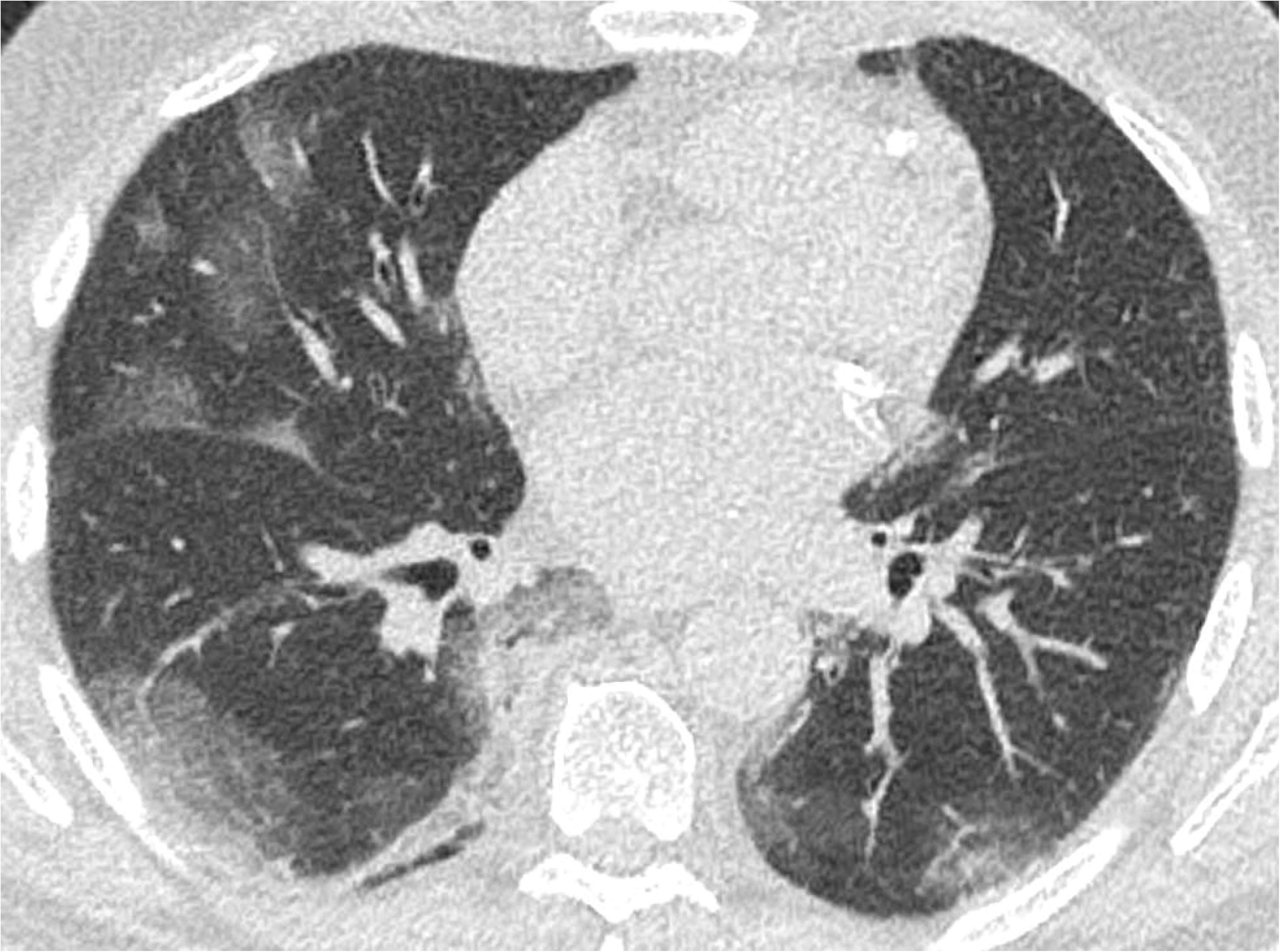
Chest CT-scan categorized « evocative of COVID-19 pneumonia » in two different patients with positive nasopharyngeal SARS-CoV-2 RT-PCR at admission and secondarly (Panels a, b and panels c, d, respectively). Multifocal bilateral ground-glass opacities with subpleural and posterior predominance, associated with band-like (panel b) or more extensive consolidations (Panel d).

**Figure 3.**
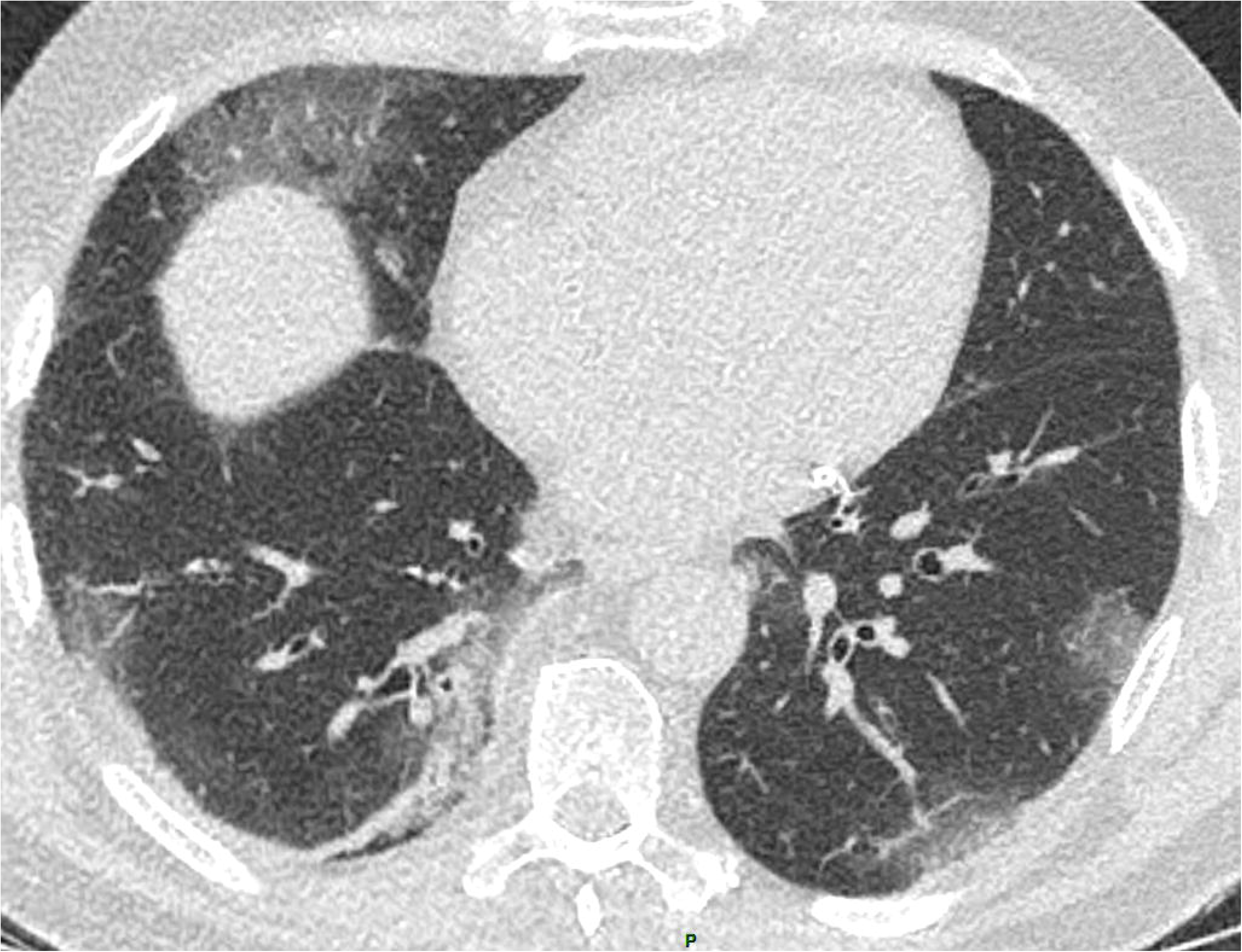
Chest CT-scan categorized « compatible with COVID-19 pneumonia », in association with fibrosing interstitial lung disease (ILD) showing a non specific interstitial pneumonia pattern (Panel c, axial plane through the lung bases). Pure ground-glass opacities, both centrally and peripherally distributed, have appeared in the left upper lobe (Panels a, b), as compared to the previous CT performed 4 months earlier (Panel d). Such new opacities could be attributable to COVID-19, another infection, or acutisation of ILD. Nasopharyngeal SARS-CoV-2 RT-PCR positive.

**Figure 4.**
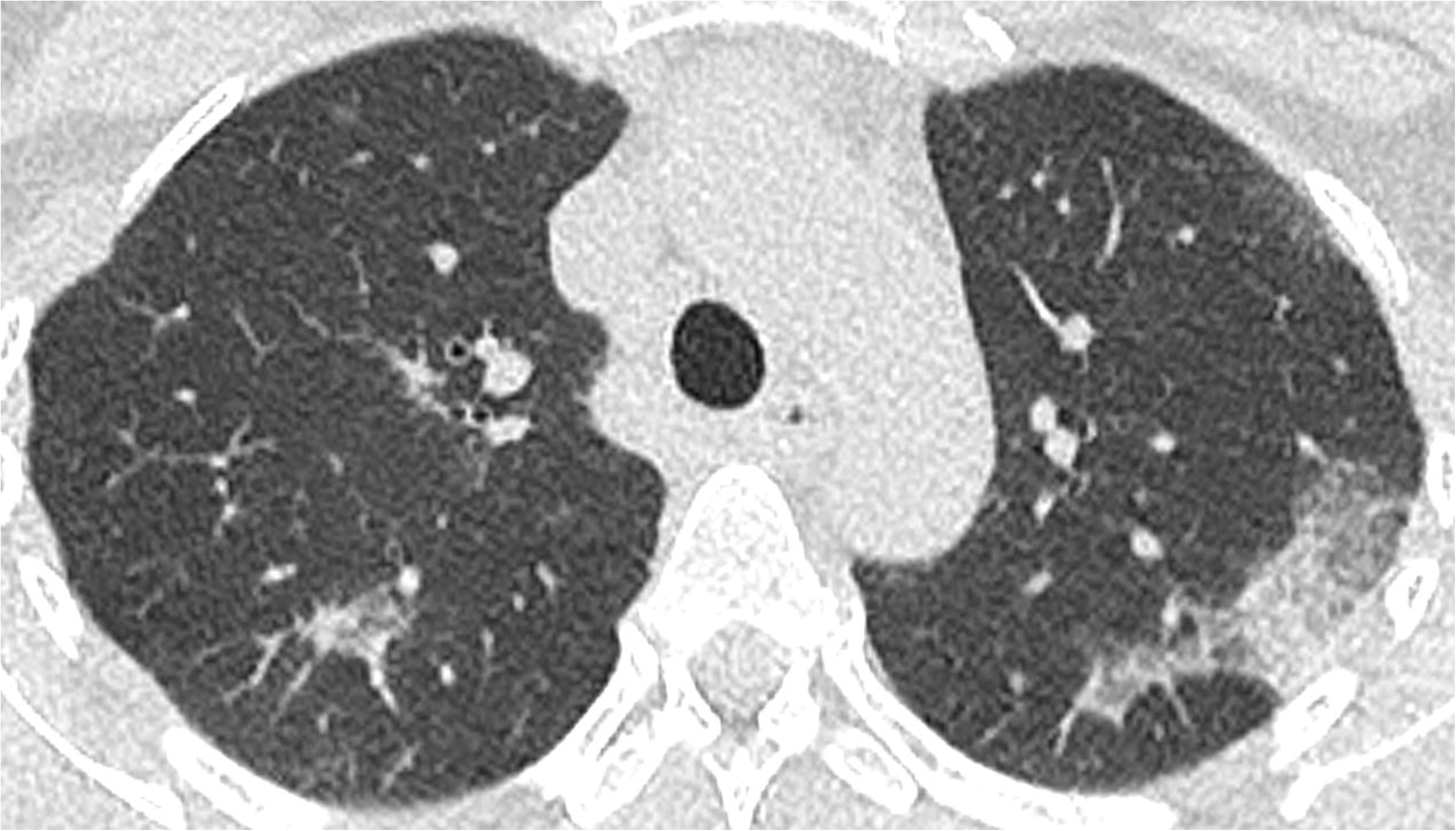
Chest CT-scan categorized « compatible with with COVID-19 pneumonia », in association with pulmonary edema, manifesting as ground-glass opacities with a predominant central distribution,septal lines and bilateral pleural effusion (Panels a, b, axial plane, Panel c, coronal plane) in a patient with history of chronic renal insufficiency on dialysis. Subpleural consolidation (Panel a) in the posterior zone of the right upper lobe is consistent with associated COVID-19 pneumonia. Nasopharyngeal SARS-CoV-2 RT-PCR positive.

**Figure 5.**
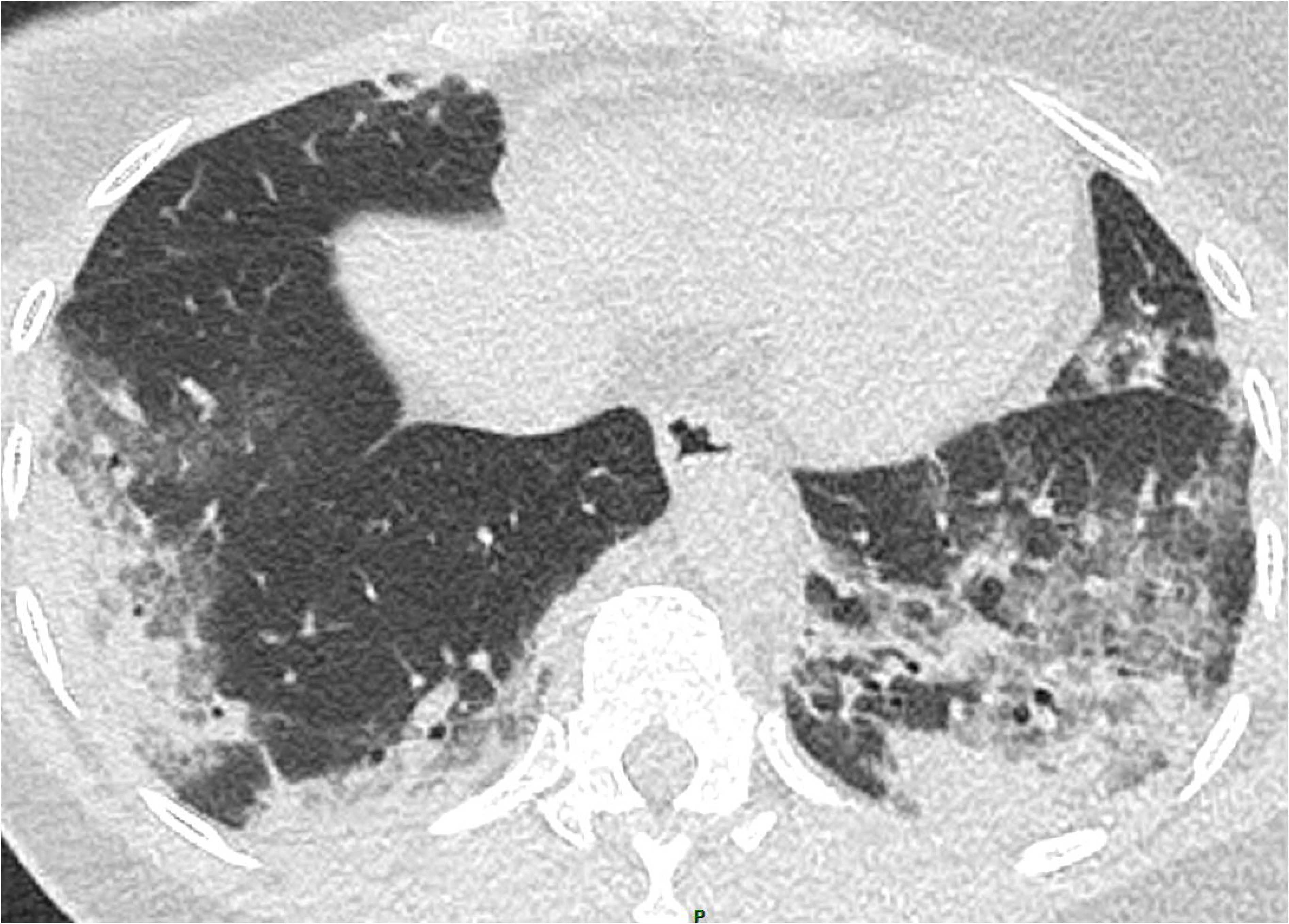
Chest CT-scan categorized « not evocative of COVID-19 peumonia », showing small consolidations in the left lower lobe associated with bronchial thickening and endobronchial filling, in favor of bronchopneumonia. Nasopharyngeal SARS-CoV-2 RT-PCR negative.

**Figure 6.**
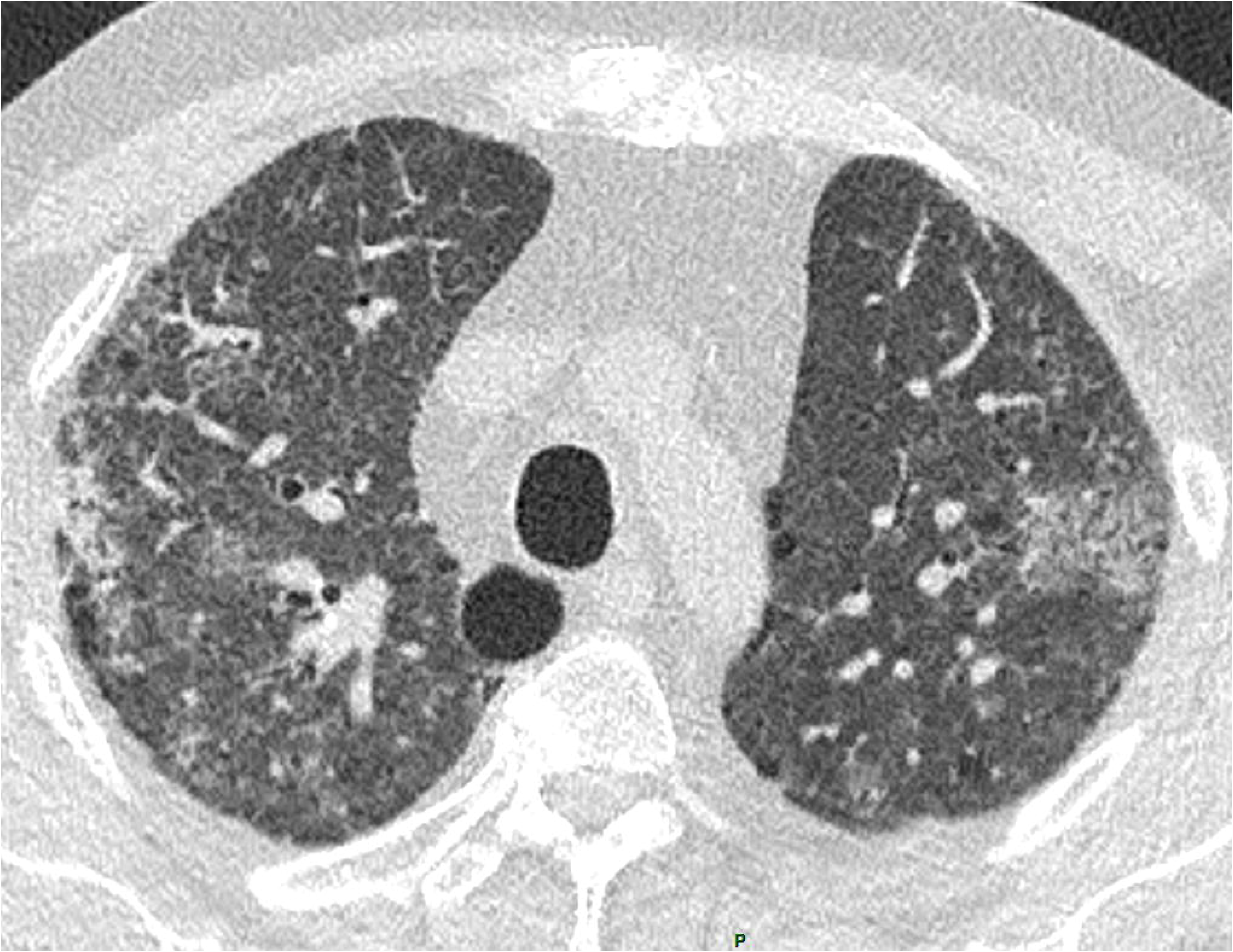
Chest CT-scan categorized « not evocative of COVID-19 pneumonia » showing combination of areas of ground-glass opacity and consolidation systematized in the middle lobe (arrows), evoquing a lobar pneumonia. Despite positivity of the SARS-CoV-2-RT-PCR, bacterial co-infection was suspected and the patient received antibiotics with favorable evolution.

## Discussion

The current study describes observer agreement for chest CT reporting in a large series of consecutive patients suspected of COVID-19. We found that categorization of CT reports was reproducible and meaningful in patients considered for hospitalization. With SARS-CoV-2 RT-PCR as reference, chest CT reported «evocative of COVID-19 pneumonia» was highly predictive of the disease in this population in the current outbreak and agreement for this category between observers of various experiences and sub-specialties was very good. The positivity rate of RT-PCR was highly significantly different among the categories, supporting a role for disease likelihood stratification by CT. It should be emphasized that a quarter of patients with chest CT classified «not evocative or normal» had a positive RT-PCR, highlighting that no CT pattern can rule out COVID-19 in the present epidemic context. As previously reported [4,17], RT-PCR may be positive in patients without lung abnormalities on CT.

We observed only fair observer’s agreement for the category «compatible». This may be explained by the method used to classify patients. Thus, categorization of chest CT was mainly based on the global impression of each reader, according to the routine practice in our hospital, where more than thousand chest CTs have been performed to date in COVID-19 patients. The guide we provided to all observers before readings to help cases classification partly relied on interpretation of findings which may vary according to each reader experience. Cases classified «compatible» encompassed only 12% of all, i. e. 30 cases at all. More than half of them showed features of an underlying pulmonary disease or of a mixed pattern with some opacities that could be attributable to pulmonary edema. Such mixed features complicate interpretation and categorization of findings, and lowers the reader’s confidence. Indeed we observed significant lower observer agreement in cases showing an underlying pulmonary disease. These mixed and complex cases are part of the routine practice.

The recent Radiological Society of North America proposal for CT findings related to COVID-19, includes 4 categories: typical, indeterminate, atypical appearance, or CT negative for pneumonia [6], the «indeterminate» category appearing similar to the «compatible» category we used in the present study. Terms and categories proposed by other Radiology Societies vary, according to whether or not they individualize a normal category, and to the intermediate category being named either «compatible» or «indeterminate» [13-15]. We herein chose to retain 3 categories, merging a normal appearance of the lung parenchyma with non-specific lung abnormalities or features suggesting an alternative diagnosis because we thought the probability of COVID-19 would be lower in these latter situations, although non-zero, and in accordance with the guidelines of the European Society of Radiology.

By showing significant differences in the RT-PCR positivity rate among CT categories, our study supports that chest CT can participate in estimating the likelihood of COVID-19, in association with contact history, clinical presentation and prevalence of the disease in the population [18]. The role of chest CT for patients suspected of COVID-19 is not completely established. Despite limitations in sensitivity and result delays, the RT-PCR remains the diagnostic reference and chest CT is not recommended for screening by most Radiology Societies [6,15,19,20]. According to a recent consensus statement from the Fleischner Society [20], imaging may be indicated for diagnosis when RT-PCR is negative or unavailable in patients having risk factors for worsening or moderate-to-severe respiratory signs. In our study, most patients who had chest CT at the emergency room had indeed either moderate or severe clinical features or comorbidities. Chest CT helped addressing or transferring patients into the proper, COVID-19 or not COVID-19, hospitalization area, especially those needing urgent decision, before the RT-PCR result was provided. Chest CT could favor re-testing in cases with negative RT-PCR [12]. Patients with a first negative RT-PCR and a chest CT considered «compatible» have been more frequently re-tested and had a subsequent PCR more frequently positive, as compared to patients with a first negative RT-PCR and a chest CT considered «not evocative». Of note, two patients had a final COVID-19 retained diagnosis based on typical clinical and CT presentation and evolution, despite two negative nasopharyngeal RT-PCR tests.

To date, performances of chest CT have been analyzed according to a binary consideration, i.e. CT positive or negative for COVID-19 pneumonia, using RT-PCR as reference. Most studies have reported high sensitivity, up to 97% [4,5,21] and specificity between 25 and 56%, with pooled sensitivity and specificity of 94% and 37%, respectively according to a recent meta-analysis [22]. Whether positive CTs in these studies showed typical imaging features is unclear. Our results differ, chest CT classified evocative having 75% sensitivity and 95% specificity and chest CT classified evocative or compatible having 85% sensitivity and 77% specificity. These differences may be attributable to differences in CT features between an «evocative» CT and a «positive» CT as well as differences in characteristics of the population having chest CT. The prevalence of the disease in the population, severity and type of clinical presentation, time from symptom to CT, age of patients and any underlying pulmonary pathology may modify the performances of chest CT for diagnosing COVID-19 pneumonia [20,23]. Indeed we observed that the presence of an underlying pulmonary disease lowered the sensitivity for an evocative CT. This concerned a quarter of the whole population in our study and almost a quarter of the patients with positive RT-PCR. CT reporting in several categories seems best suited to the routine practice than a binary conclusion, when CT abnormalities may mix different types of lesions or are very limited in extent. It allows identifying a category with typical features, whose high specificity can be useful in an epidemic context, allowing relying on chest CT for diagnosis in some cases. Our study has some limitations. Firstly, because it is monocentric and because of various presentation and prevalence of the disease around the world, caution should be taken to extrapolate CT sensitivity and specificity between populations and periods [24,25]. We may assume that the high specificity we report for the category «evocative» would be lower if the prevalence of COVID-19 decreased and the one of other viral pneumonia or some interstitial lung diseases, as drugs or connective tissue diseases related, increased. Secondly, the clinical significance of CT reporting should integrate the risk level of each patient, that we have not precisely taken into account, even if the study period took place during the outbreak.

In conclusion, inter-observer agreement to report chest CT findings into categories for clinical suspicion of COVID-19 is good, among readers of various experience levels and sub-specialties. Chest CT can participate in estimating the likelihood of COVID-19 in patients presenting to hospital during the outbreak. CT categorized evocative of COVID-19 pneumonia were highly predictive of the disease, whereas the predictive value of CT decreased between the categories «compatible» and «not evocative or normal», from 50 to 27%. Category reports need to be integrated to the clinical presentation and risk level for COVID-19.

## Abbreviations

COVID-19: Coronavirus disease 2019

SARS-CoV-2: severe acute respiratory syndrome coronavirus 2

RT-PCR: Real-time reverse transcription-polymerase chain reaction

CT: Computed tomography

GGO: Ground-glass opacities

## Data Availability

all data referred to in the manuscript are available (email address of the corresponding author)

---

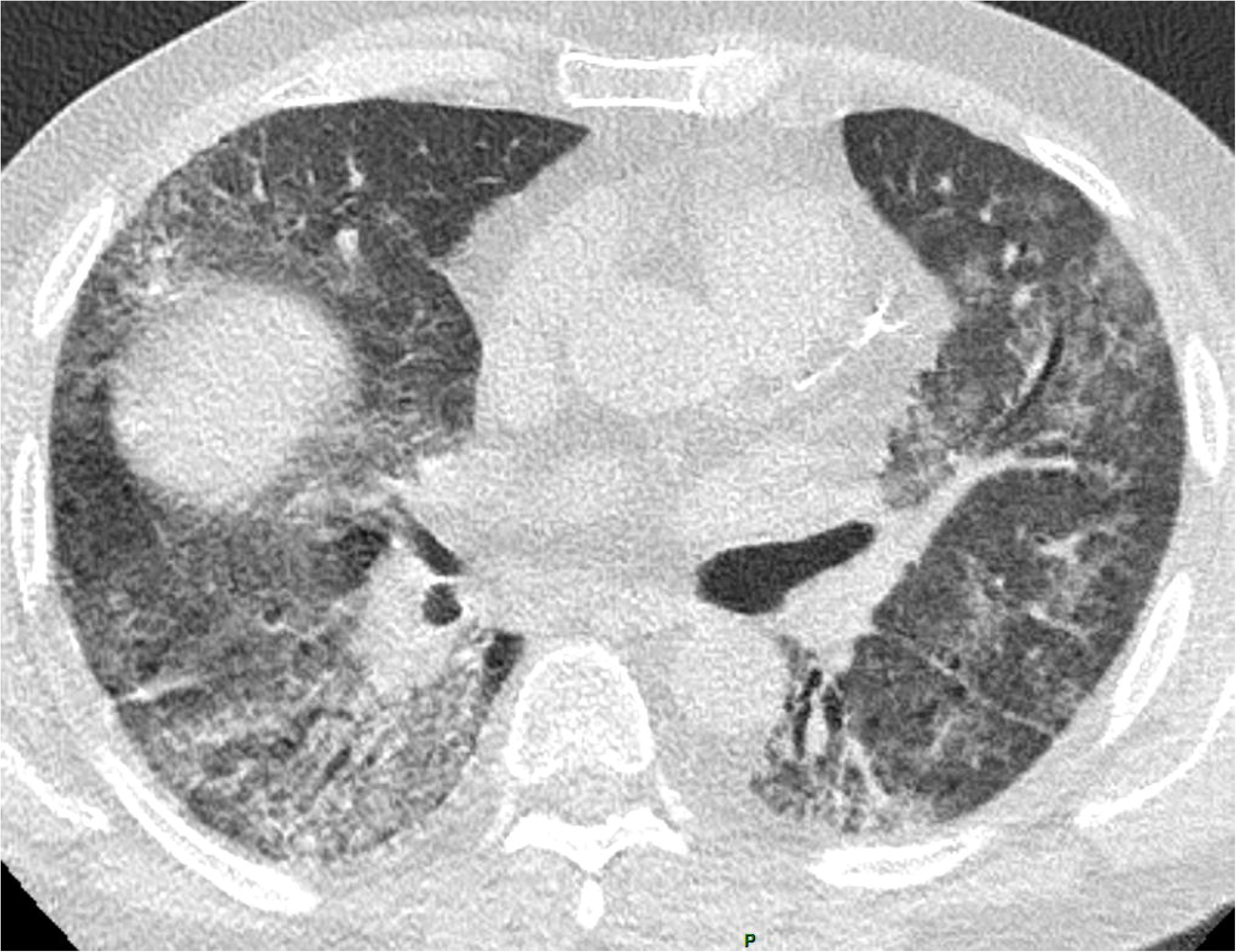

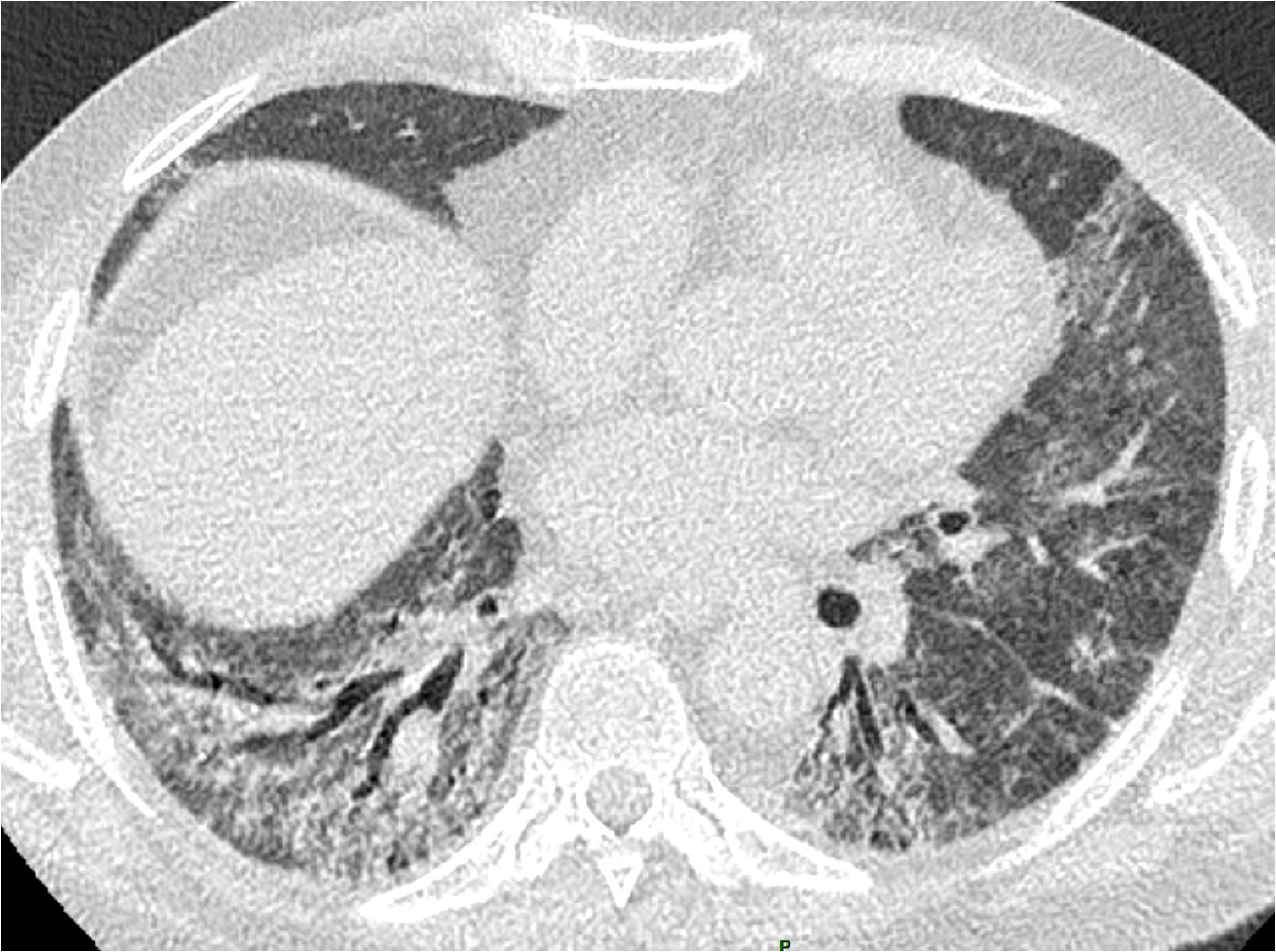

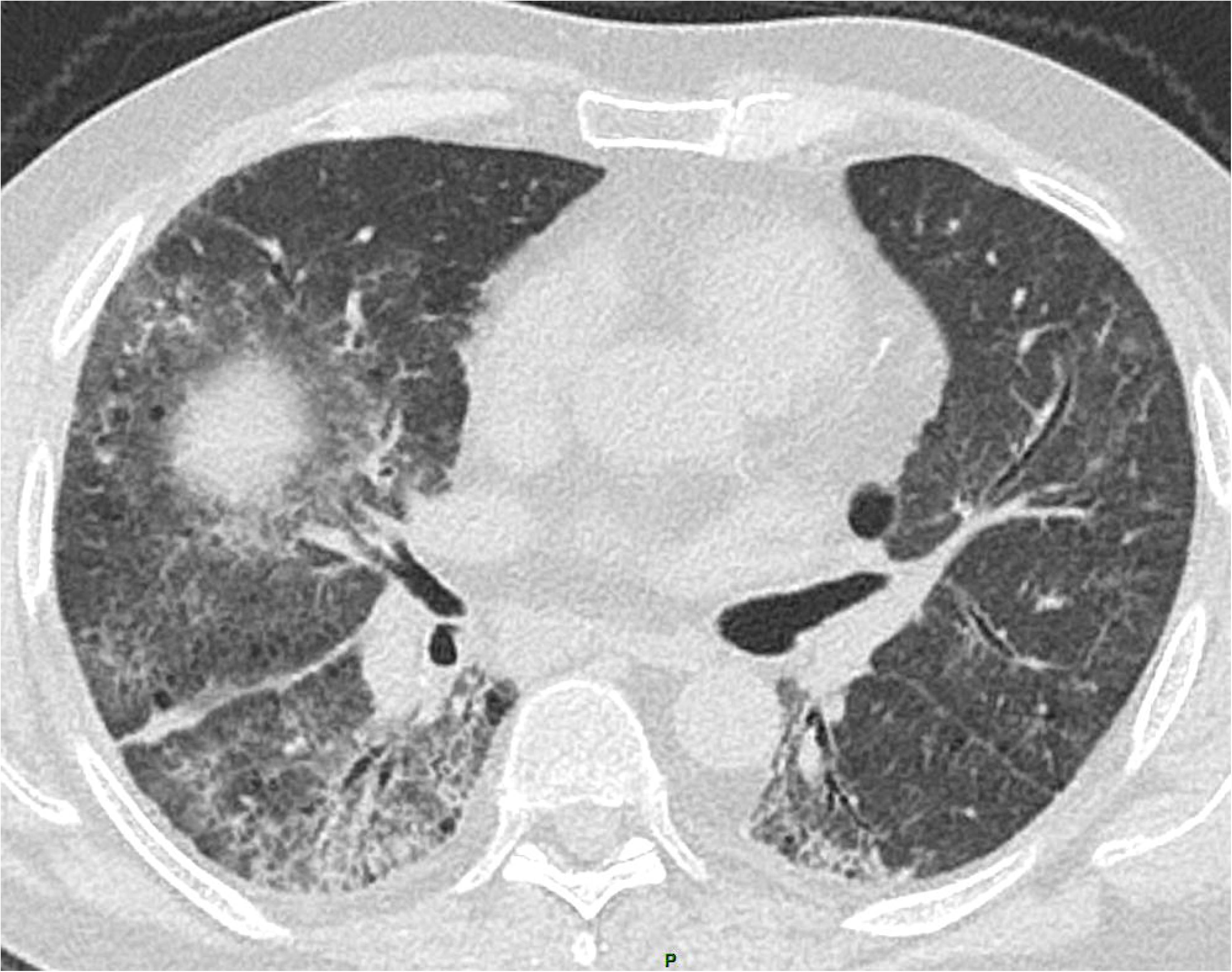

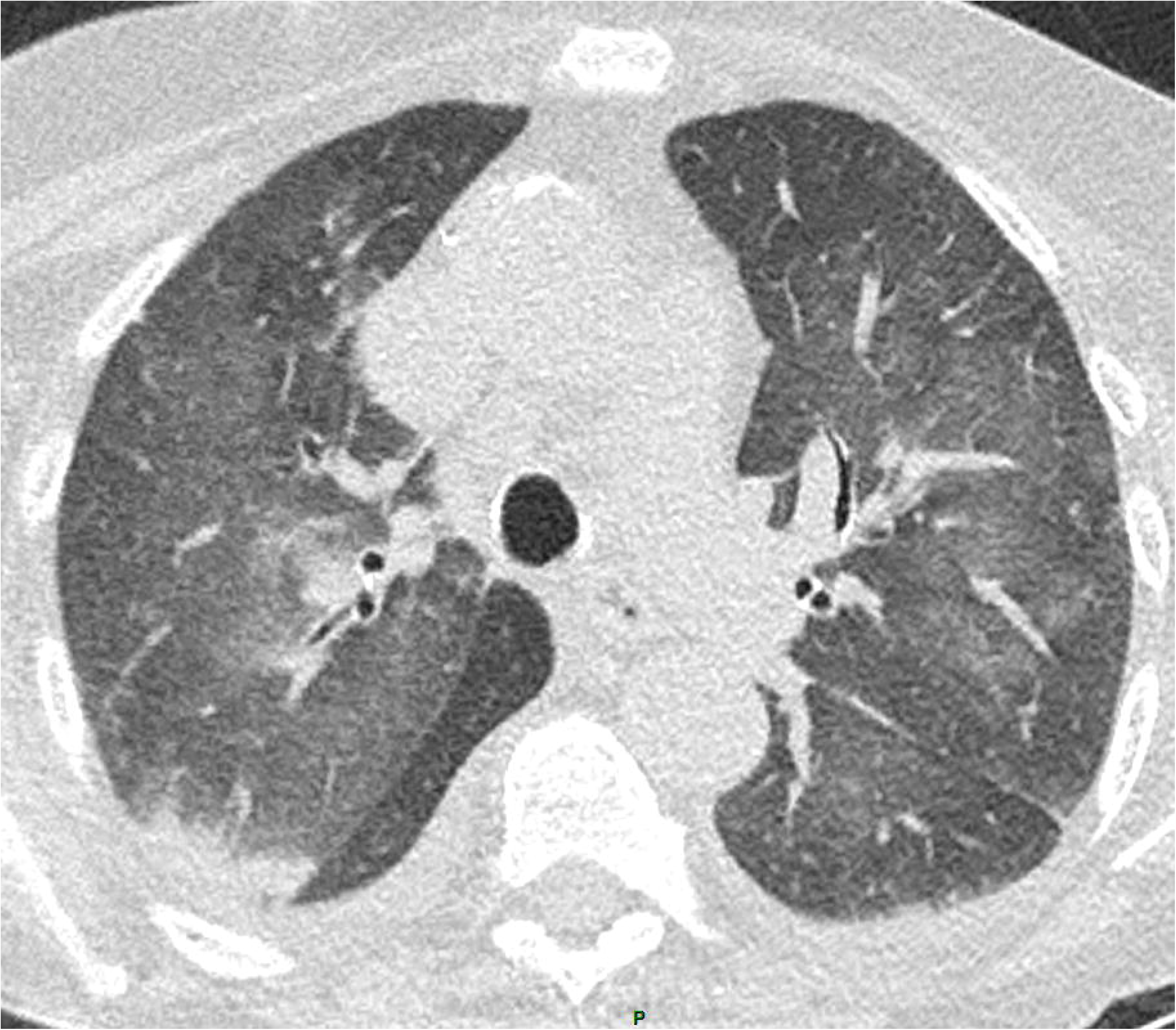

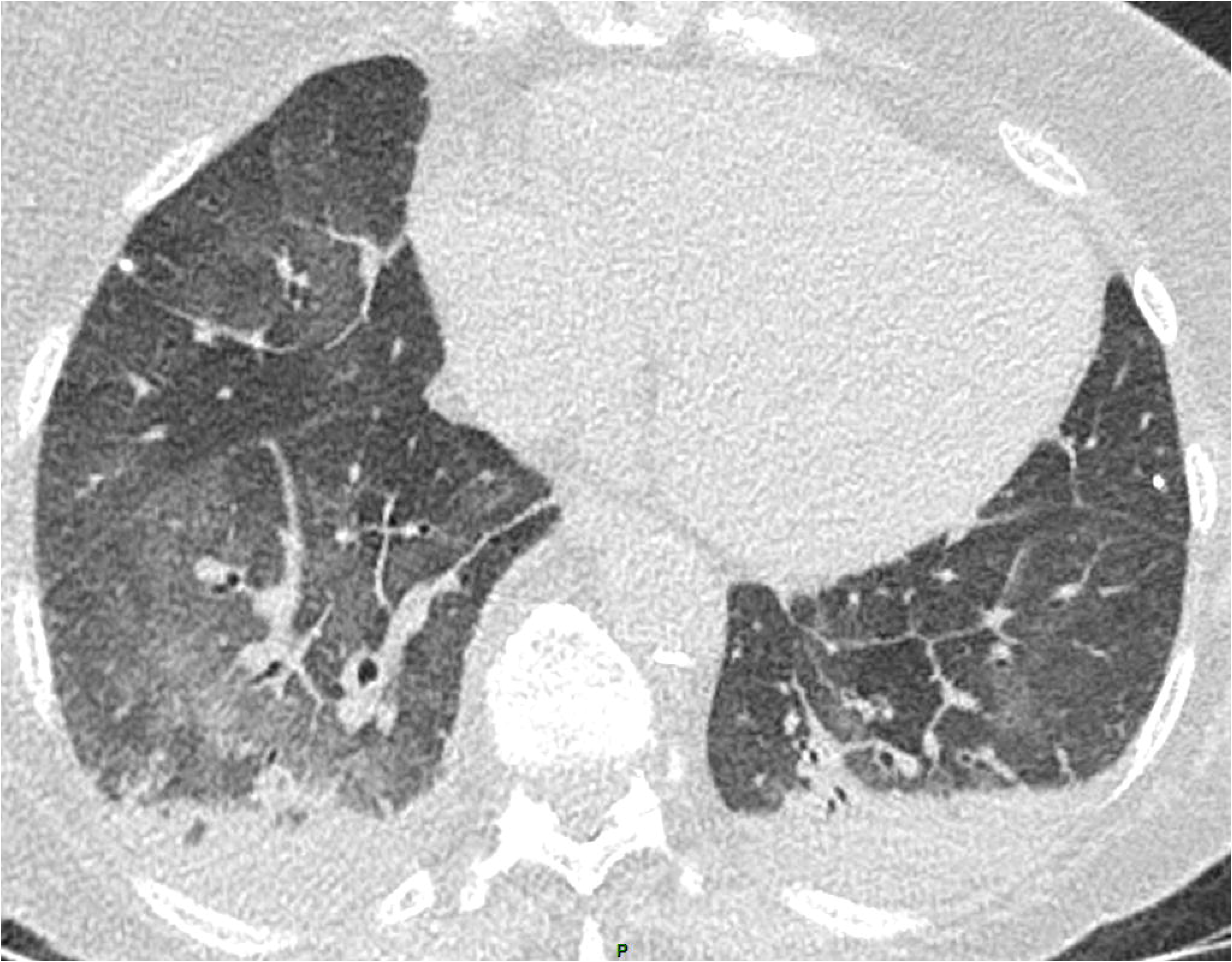

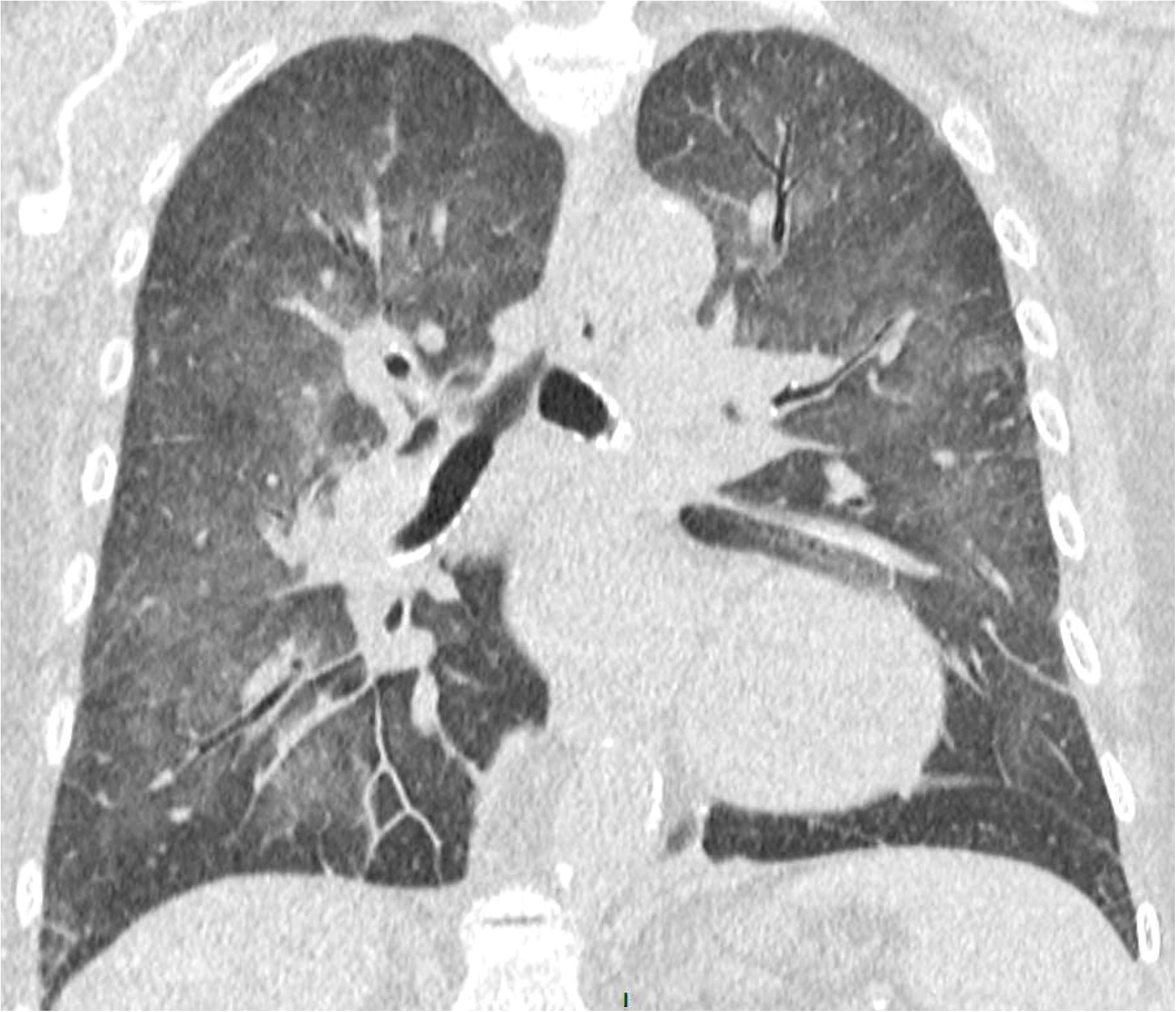

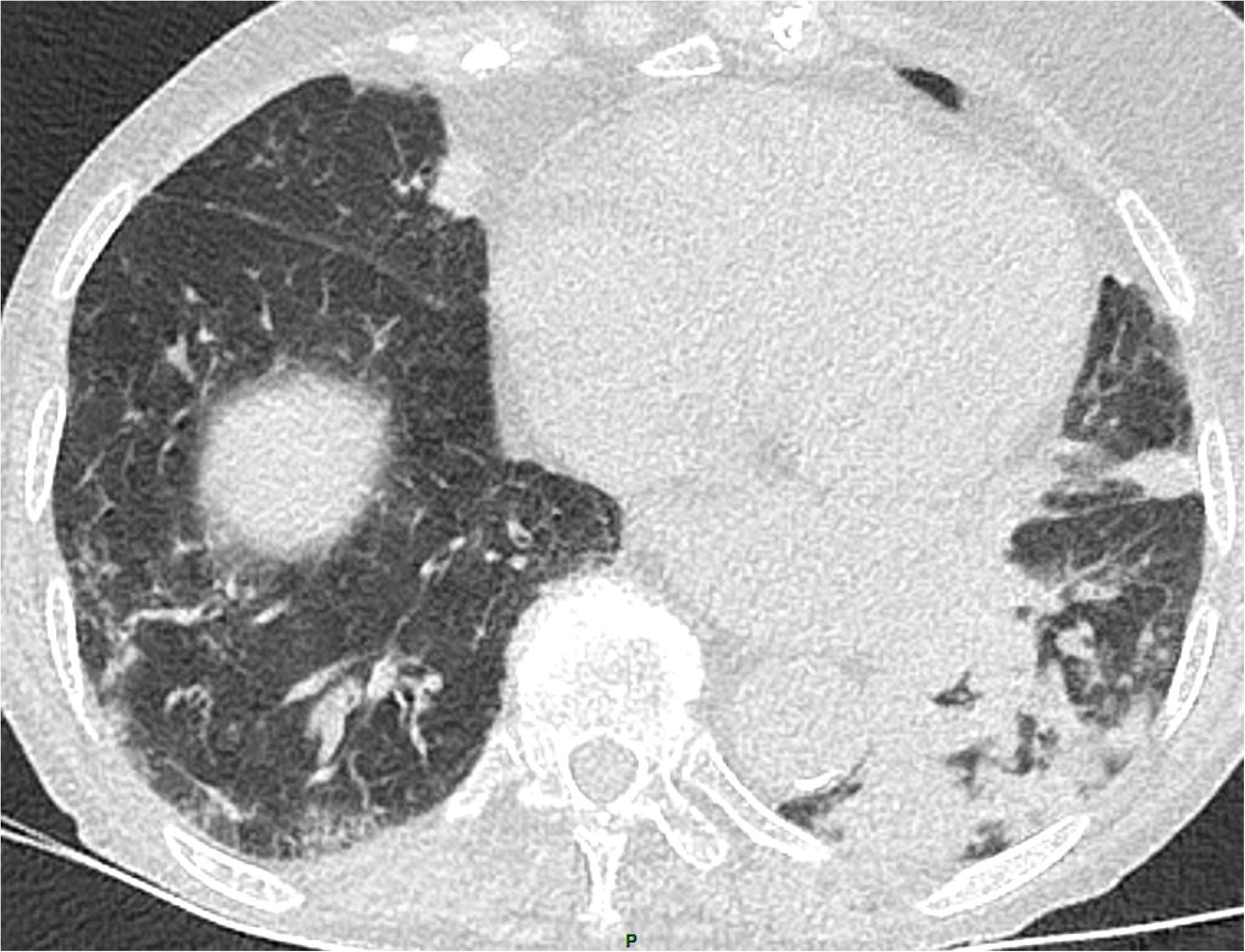

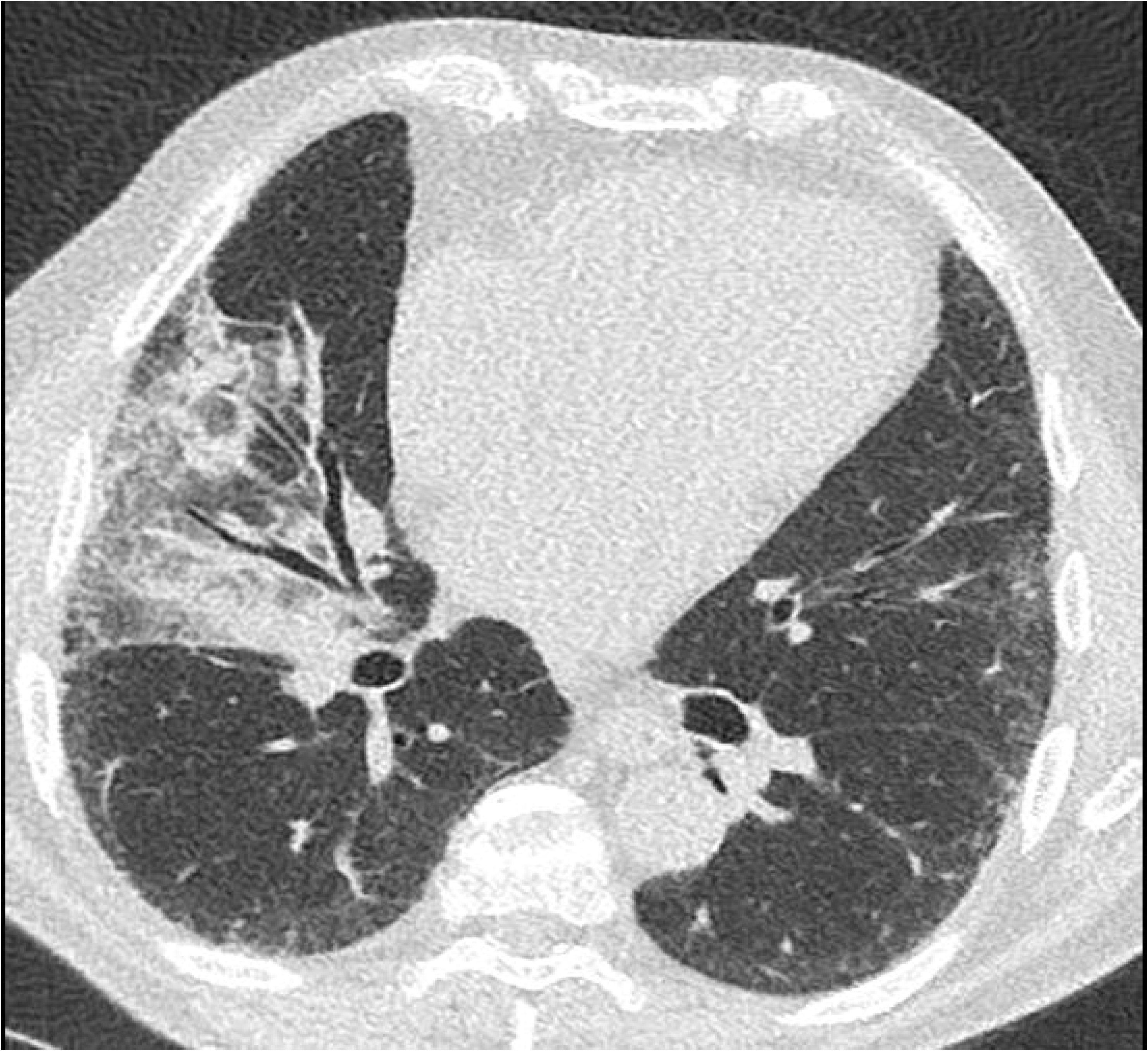

